# Inhibitor screening of Spike variants reveals the heterogeneity of neutralizing antibodies to COVID-19 infection and vaccination

**DOI:** 10.1101/2021.05.15.21257254

**Authors:** Xiaomei Zhang, Mei Zheng, Te Liang, Haijian Zhou, Hongye Wang, Jiahui Zhang, Jing Ren, Huoying Peng, Siping Li, Haodong Bian, Chundi Wei, Shangqi Yin, Chaonan He, Ying Han, Minghui Li, Xuexin Hou, Jie Zhang, Liangzhi Xie, Jing Lv, Biao Kan, Yajie Wang, Xiaobo Yu

## Abstract

Mutations of the coronavirus responsible for coronavirus disease 2019 (COVID-19) could impede drug development and reduce the efficacy of COVID-19 vaccines. Here, we developed a multiplexed Spike-ACE2 Inhibitor Screening (mSAIS) assay that can measure the neutralizing effect of antibodies across numerous variants of the coronavirus’s Spike (S) protein simultaneously. By screening purified antibodies and serum from convalescent COVID-19 patients and vaccinees against 72 S variants with the mSAIS assay, we identified new S mutations that are sensitive and resistant to neutralization. Serum from both infected and vaccinated groups with a high titer of neutralizing antibodies (NAbs) displayed a broader capacity to neutralize S variants than serum with low titer NAbs. These data were validated using serum from a large vaccinated cohort (n=104) with a tiled S peptide microarray. In addition, similar results were obtained using a SARS-CoV-2 pseudovirus neutralization assay specific for wild-type S and four prevalent S variants (D614G, B.1.1.7, B.1.351, P.1), thus demonstrating that high antibody diversity is associated with high NAb titers. Our results demonstrate the utility of the mSAIS platform in screening NAbs. Moreover, we show that heterogeneous antibody populations provide a more protective effect against S variants, which may help direct COVID-19 vaccine and drug development.

**Highlights:**

- Developed a high throughput assay to screen the neutralizing effect of antibodies across multiple SARS-CoV-2 Spike variants simultaneously.
- Characterized the heterogeneity of neutralizing antibodies produced in response to COVID-19 infection and vaccination.
- Demonstrated the capacity of Spike variants neutralization is associated with the diversity of anti-Spike antibodies.

## Introduction

Coronavirus disease 2019 (COVID-19) is caused by the severe respiratory coronavirus 2 (SARS-CoV-2). As of May 2021, SARS-CoV-2 had infected 162 million people and caused over three million deaths worldwide (Dong et al., 2020). A key step in infection is viral entry, which is facilitated by the interaction between the SARS-CoV-2 Spike (S) protein via its receptor binding domain (RBD) (319 – 541 aa) with the human Angiotensin-Converting Enzyme 2 (ACE2) receptor. Thus, this interaction is a major focus in drug and vaccine development efforts (Burki, 2021; Dai and Gao, 2021). For example, the current COVID-19 vaccines approved by the U.S. Food and Drug Administration (FDA) stimulate the immune response to generate antibodies capable of neutralizing the Spike-ACE2 interaction. Unfortunately, SARS-CoV-2 is mutating, with new variants emerging nearly every week that could impede drug development and reduce the efficacy of COVID-19 vaccines (Dai and Gao, 2021; Kemp et al., 2021).

Mutations in the S protein are of particular concern since they could enable SARS-CoV-2 to evade defense mechanisms that are elicited by COVID-19 vaccines and therapeutic antibodies (Aschwanden, 2021; Del Rio and Malani, 2021). For example, the D614G variant, which was first identified in July 2020, has a faster infection rate and higher viral load in the upper respiratory tract than the wild-type “Wuhan-Hu-1” strain (Hou et al., 2020b; Korber et al., 2020). It has since become one the most prevalent strains. The B.1.1.7 variant (D614G, N501Y) is more infectious and may lead to increased mortality compared to the parental strain (Davies et al., 2021; Galloway et al., 2021). The B.1.351 and P.1 variants contain three RBD mutations at E484K, N501Y, and K417N or K417T, respectively. These mutations have shown resistance to neutralizing antibodies (NAbs) produced by convalescent COVID-19 patients and vaccinees that inhibit the wild-type Spike-ACE2 interaction (Garcia-Beltran et al., 2021; Liu et al., 2021; Madhi et al., 2021; Wang et al., 2021a; Wang et al., 2021b; Wibmer et al., 2021). The vaccinees in these studies received the most popular vaccines worldwide, including mRNA-based COVID-19 vaccines (Moderna and Pfizer BioNTech) and a replication-deficient chimpanzee adenoviral vector COVID-19 vaccine (AstraZeneca). Similar results were obtained when testing a B.1.1.7 variant with an additional E484K mutation (Collier et al., 2021; Wang et al., 2021b). These studies highlight the importance of an assay that can measure the humoral response to S variants in developing effective therapeutic antibodies and vaccines for COVID-19 (Burki, 2021; Priesemann et al., 2021).

To address this urgent need, we developed a protein microarray for the high throughput, multiplexed detection of NAbs to SARS-CoV-2 S variants. This multiplexed Spike-ACE2 Inhibitor Screening (mSAIS) assay is simple to use, able to detect NAbs to numerous S variants simultaneously, requires minimal sample volume (i.e., 20 µL serum), and can be performed with common laboratory equipment. It could also be used with other potential neutralizing molecules (e.g., small molecules). To demonstrate the potential of the mSAIS assay, we assessed the neutralization potential of purified anti-S antibodies and serum from convalescent COVID-19 patients and vaccinees across 72 S variants. The sensitivity and resistance of the various S protein mutations to the NAbs were determined, and new escape mutations that are not targeted by vaccine-induced antibodies were identified. The neutralization capacity of high and low titer NAb titer samples was also compared. Our results were validated using a peptide-based microarray and SARS-CoV-2 pseudovirus neutralization assay.

## Results

### Development of the mSAIS assay

The mSAIS assay enables the simultaneous screening of various potential neutralizing molecules across numerous SARS-CoV-2 S variants within 2 hours (Figure 1A). Here, 72 S protein variants with a polyhistidine tag at the C-terminus were expressed in the human embryonic kidney 293 (HEK293) cell line (Figure S1), purified, and printed onto a chemically-modified glass slide as previously described (Wang et al., 2020; Zhang et al., 2020b). The printed S protein variants were selected from the COVID-19 virus mutation tracker database (Figure S2) and literature, and included 2 variants with mutations in the S protein’s subunit 1 (S1) and subunit 2 (S2) domains, 7 variants with mutations in S1, and 63 variants with mutations in the RBD (Alam et al., 2021). Prevalent SARS-CoV-2 strains, such as the D614G, B.1.1.7, and B.1.351 strains, were among the variants printed. In addition, two negative controls and one positive control (SARS-CoV-2 wild-type RBD) were printed on the array (Figure S3). The negative controls included on the array were the SARS-CoV-2 Nucleocapsid (N) protein and the RBD of the Middle East respiratory syndrome coronavirus 2 (MERS-CoV-2). Following printing, the sample (e.g., NAb) and a Cy5-labeled extracellular domain of ACE2 were sequentially incubated on the array with alternated wash steps to remove unbound sample and ACE2. Thus, S-ACE2 complex formation resulted in fluorescence whereas neutralizing samples decreased fluorescence.

**Figure 1.**
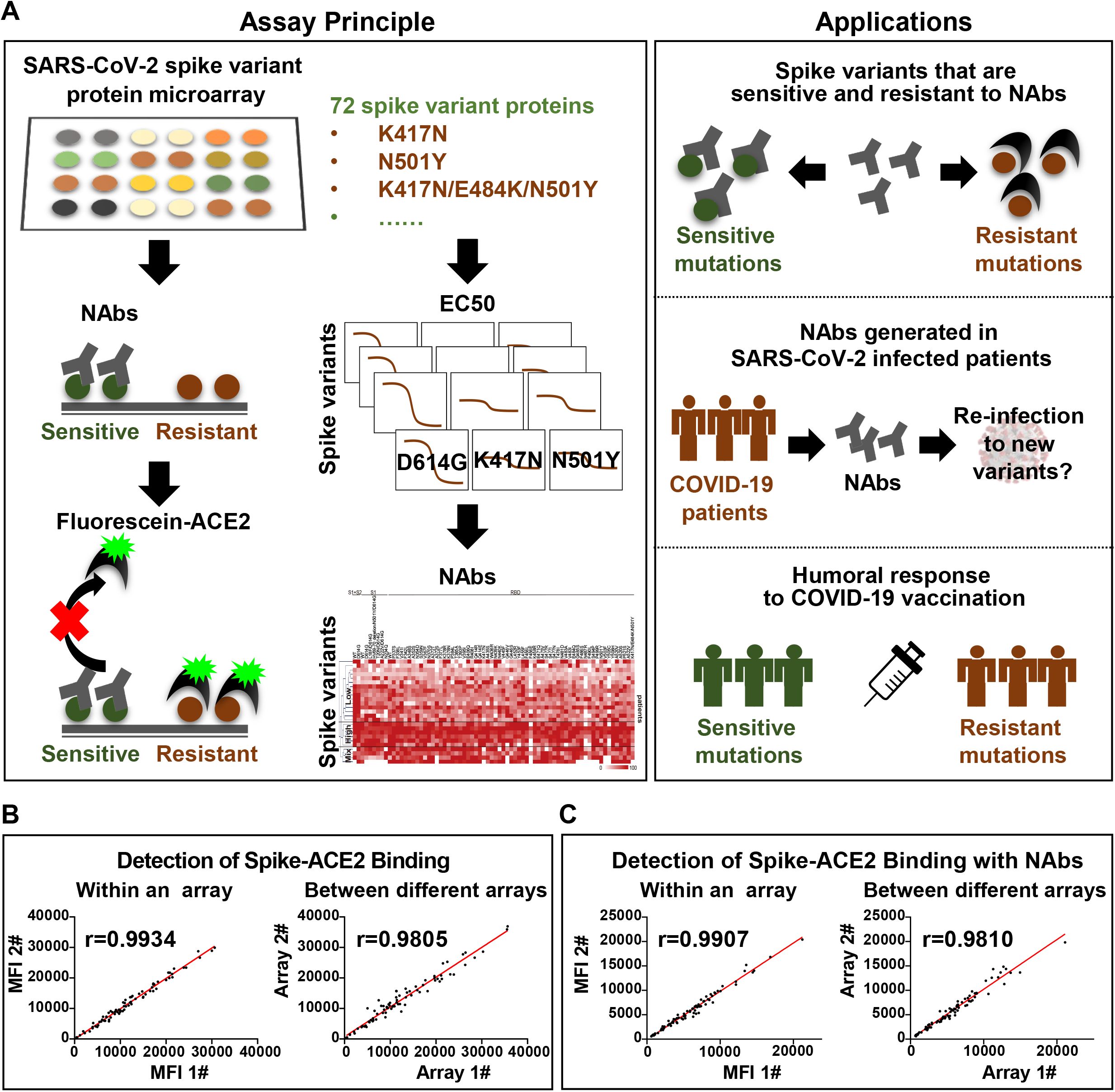
Development of the mSAIS assay. (A) A schematic illustration of the microarray-based mSAIS assay and potential biomedical applications; (B, C) Intra- and inter-array reproducibility of the mSAIS assay in the absence and presence of neutralizing antibodies, respectively.

Before using the mSAIS assay to screen samples, the reproducibility of the array and the binding of the S variants with ACE2 in the presence and absence of NAbs were evaluated. The results show that the intra- and inter-array *r* correlations for each step of the assay were 0.99 and 0.98, respectively (Figure S4, Figure 1B and 1C). Furthermore, the neutralizing capacity of 13 serum samples from vaccinees determined by the mSAIS assay was compared to the data obtained using the live SARS-CoV-2. The *r* correlation between the two approaches was 0.82 (Figure S5), demonstrating that the mSAIS assay has high reproducibility and is a feasible approach for screening potential neutralizing molecules. It is also important to note that working with live SARS-CoV-2 requires biosafety level 3 (BSL3) facilities whereas the mSAIS assay can be performed safely at BSL1 or BSL2 depending on the nature of the tested samples.

### Characterizing ACE2 interactions with Spike variants with the mSAIS assay

By testing different amounts of purified antibodies or serum, the half maximal inhibitory concentration (IC50) or effective concentration (EC50) can be determined, respectively, and the results visualized via fluorescence. When the concentration of ACE2 applied to the assay was low (≤ 2.5 µg/mL), the fluorescent signal across the different S variants varied, thus indicating that the binding affinities of ACE2 to the S variants are not the same (Figure 2A). Indeed, the calculation of the EC50 for the S variants with mutations between residues 234 - 614 shows that the binding affinity (EC50) ranges from 0.65 to 17.25 µg/mL. ACE2 had the lowest binding affinity to two S variants, F465E and N487R, with an EC50 of 16.16 and 17.25 µg/mL, respectively (Figure 2B). The EC50 ratios between the wild-type and variant S proteins were calculated and log2 transformed (Figure 2C). Using a 2-fold minimum as the selection criteria, 10 mutations that weaken the S-ACE2 interaction were identified, including A372T, F377L, G446V, F456E, G485S, F486S, N487R, F490L, P499R and Y505C (Figure 2C and 2D). Notably, the S variant D614G did not appear to affect ACE2 binding, which agrees with a previous study using surface plasmon resonance (Zhang et al., 2020a).

**Figure 2.**
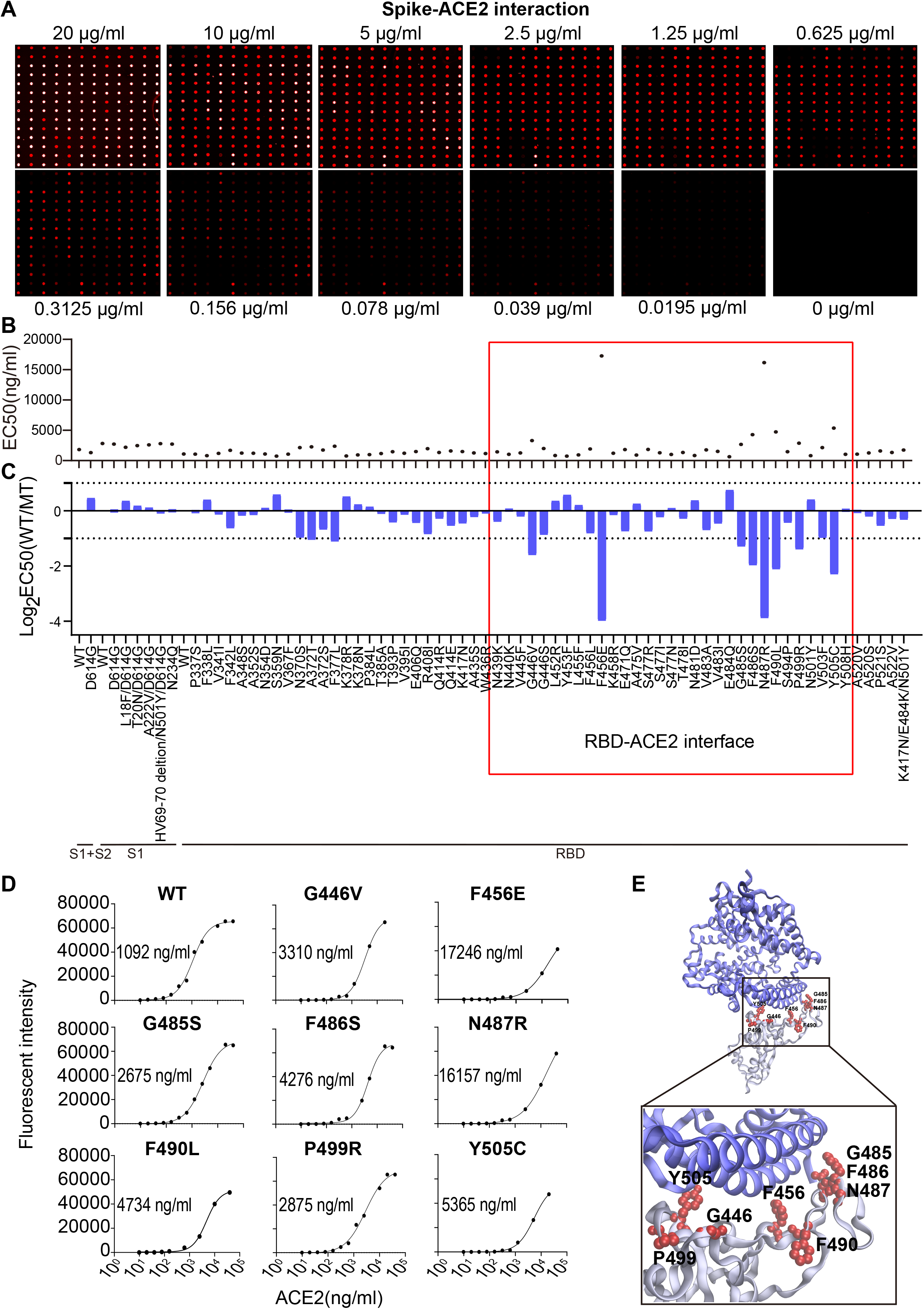
SARS-CoV-2 S-ACE2 interaction mapping across different S variants using the mSAIS assay. (A) Detection of different concentrations of ACE2 binding to S variants immobilized on the array. (B) Distribution of S-ACE2 binding affinities (EC50) across different SARS-CoV-2 S variants. (C) Fold changes of binding affinity between the wild-type and mutated S proteins. The x-axis represents the S variants and the y-axis represents the log2 transformed IC50 ratio between the wild-type and mutated S proteins. (D) are the dynamic curves representing S variants that bind to ACE2 with significantly decreased binding affinity compared to the wild-type S protein. (E) Structural analysis of escape mutations on the RBD (gray color) that interacts with ACE2 (green color). RBD mutations (PDB ID: 6M0J) are labelled in red.

Interestingly, 8/10 (80%) mutations that decreased the ability of the S protein to bind ACE2 (G446V, F456E, G485S, F486S, N487R, F490L, P499R, Y505C) are located at the RBD-ACE2 interaction interface (Figure 2E). Moreover, F456E and N487R had the weakest ACE2 binding affinities. These results further support the importance of the RBD domain in antibody neutralization (Barnes et al., 2020; Dai and Gao, 2021; Tan et al., 2020).

### Heterogeneous reaction of antibodies to Spike variants

Three antibodies that bind to the S protein’s RBD (Figure S6) were tested with the mSAIS assay, including a mouse monoclonal antibody #73, a rabbit polyclonal antibody #21, and a rabbit monoclonal antibody #53 (Figure 3). The IC50 of antibodies #21 and #53 to the S variants range from 42.04 – 10,000 ng/mL and 0.00063 – 10,000 ng/mL, respectively (Figure 3B and 3C). However, antibody #73 did not inhibit the RBD-ACE2 interaction (Figure 3A), indicating that not all anti-RBD antibodies have neutralizing activity.

**Figure 3.**
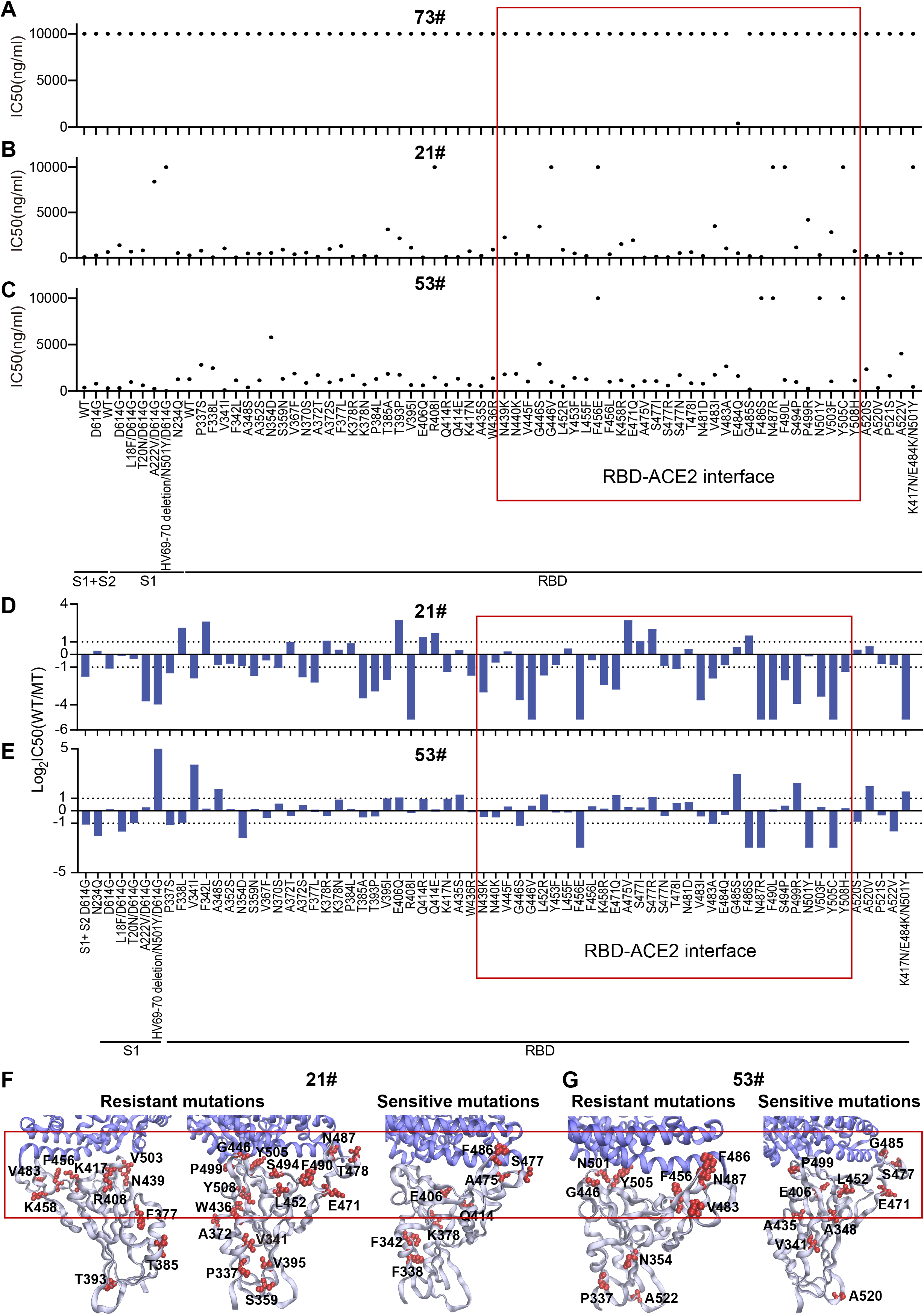
Heterogeneous responses of purified antibodies to SARS-CoV-2 mutations. (A-C) Distribution of neutralization activity (IC50) of purified antibodies #73, #21 and #53 to the S variants, respectively. The x-axis represents the S variants and the y-axis represents the neutralization activity (IC50) of the monoclonal antibody to each spike variant. (D, E) Fold changes of the neutralization activity (IC50) of antibodies #21 and #53 between the wild-type and mutant S proteins, respectively. The x-axis represents the S variants and the y-axis represents the log2 transformed IC50 ratio between the wild-type and mutated S proteins. (F, G) Structural analysis of the mutations that decrease and increase the neutralization activity of antibodies #21 and #53, respectively.

Antibody #21 was unable to neutralize the formation of the S-ACE2 complex for 8 “resistant” S variants: R408I, HV69-70 deletion/N501Y/D614G (B.1.1.7 strain), G446V, F456E, N487R, F490L, Y505C, and K417N/E484K/N501Y (B.1.351 strain) (Figure 3B). Two other studies also observed similar results for the B.1.351 strain (Li et al., 2021; Wang et al., 2021a). Using a two-fold difference in signal compared to the wild-type S protein as the criteria, 33 variants showed resistance to neutralization with antibody #21(Figure 3D). Seventeen of the 33 (51.5%) mutations are located between residues 437 and 508, which is the region of the RBD that physically interacts with ACE2 (Singh et al., 2021) (Figure 3D and 3F, Figure S7A). Four of the ten (40%) sensitive mutations are also located within the RBD interface (Figure 3D and 3F, Figure S7A).

Antibody #53 had 13 escape mutations of varying resistance, with 5 mutations (F456E, F486S, N487R, N501Y, Y505C) showing complete resistance to neutralizing activity (Figure 3C and 3E). Seven (53.85%) mutations are located within the interface of the RBD-ACE2 interaction (Figure 3E and 3G, Figure S7B). Interestingly, six (46.15%) sensitive mutations are not located within the RBD-ACE2 interface (Figure 3E and 3G, Figure S7B), which indicates that binding to non-RBD binding epitopes can also have neutralization effects (Li et al., 2020; Prevost and Finzi, 2021).

### Heterogeneous reaction of serological NAbs to S variants in convalescent COVID-19 patients

We next screened the serum from 25 COVID-19 patients who had recovered from SARS-CoV-2 infection. Using hierarchical cluster analysis, the convalescent patients clustered into three groups based on their NAb titers: high, low, and mixed (Figure 4A and 4B). After determining the percentage of escape mutations under three different EC50 thresholds (10, 30, 50), it was observed that the number of escape mutations were fewer for the high-titer NAb group than the low-titer NAb group (Figure 4C).

**Figure 4.**
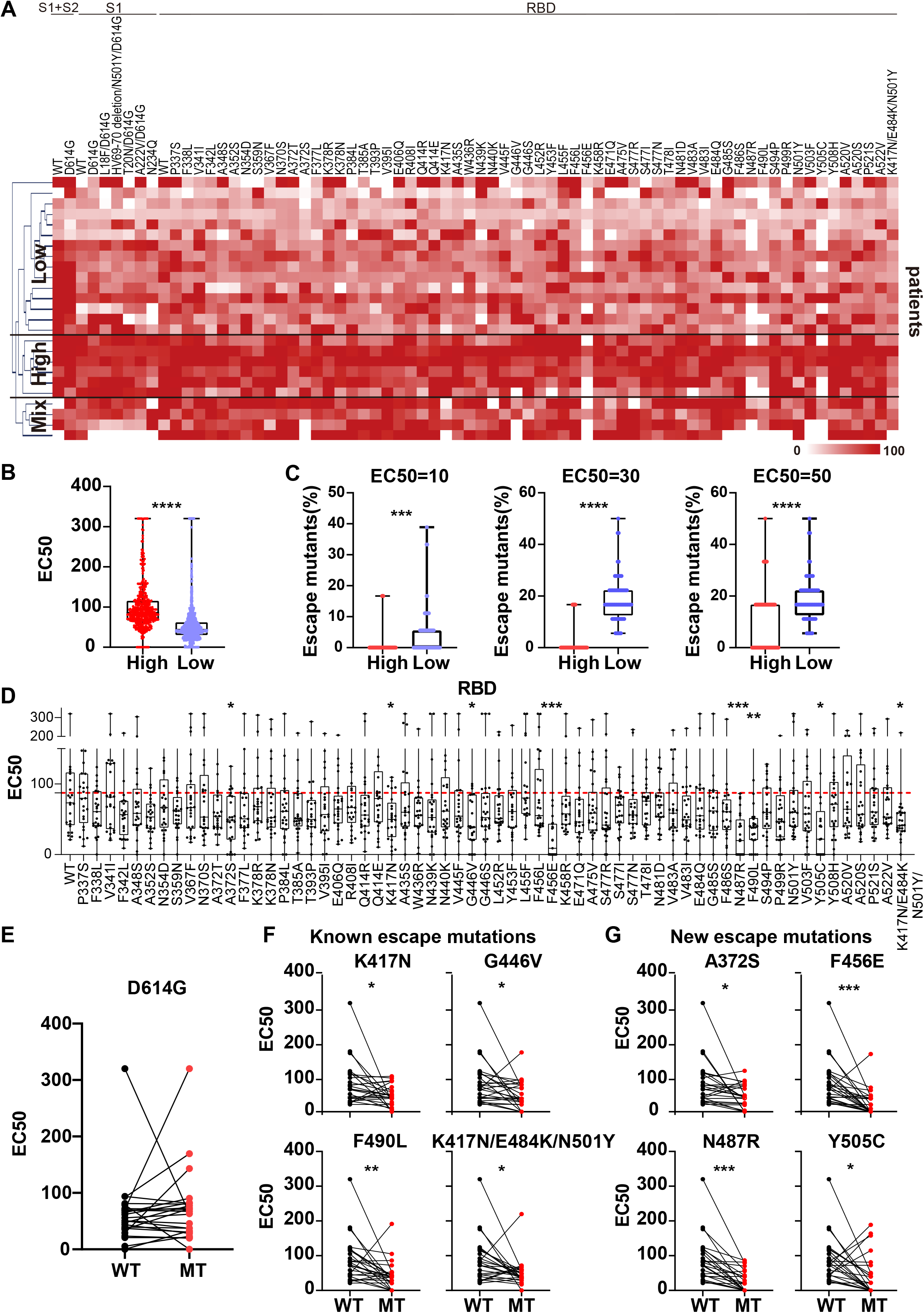
Heterogeneous responses of serological NAbs to SARS-CoV-2 S mutations in convalescent COVID-19 patients. (A) Hierarchical cluster analysis of NAbs to S variants in convalescent COVID-19 patients. The rainbow color from white to red correspond to the NAb titer from low to high, respectively. (B) EC50 comparison of convalescent COVID-19 patients with high and low NAb titers. (C) Comparison of escape mutations between convalescent COVID-19 patients with high and low NAb titers with EC50 thresholds set at 10, 30 and 50, respectively. (D) Distribution of serological NAbs to different SARS-CoV-2 S variants in convalescent COVID-19 patients. (E) Comparison of NAbs between the wild-type and D614G S proteins. (F) Comparison of NAbs between the wild-type S protein and S variants with known escape mutations. (G) Comparison of NAbs between the wild-type S protein and S variants with newly identified escape mutations. Significant escape mutations were identified using an unpaired t test with a p value of 0.05.

Next, the NAbs to wild-type and mutant S proteins were compared (Figure 4D). The D614G variant did not lead to a change of NAbs to S1 and S1+S2 proteins (Figure 4E, Figure S8). Using statistical analysis, the number of NAbs to eight mutated proteins was significantly decreased, including 4 known escape mutations [K417N, G446V, F490L, K417N/E484K/N501Y (B.1.351 strain)] and 4 newly identified escape mutations (A372S, F456E, N487R, Y505C) (Figure 4F and 4G) (Chen et al., 2021; Li et al., 2021; Li et al., 2020; Zhou et al., 2021). Notably, structural analyses show that 5 escape mutations (K417N/E484K/N501Y, G446V, F456E, N487R, F490L) are located within the interface of RBD-ACE2 interaction, thus indicating that this RBD subdomain is important in neutralizing SARS-CoV-2 infection (Wang et al., 2020). All these results demonstrate the heterogeneous reactivity of NAbs produced by COVID-19 patients across S variants.

### Heterogeneous reactivity of NAbs produced by vaccinees to S variants

The mSAIS assay was next used to measure the neutralizing effect of antibodies produced by 30 vaccinees who received the inactivated vaccine in China (Wu et al., 2021). NAbs were generated to the majority of S variants (Figure 5A). Using hierarchical cluster analysis, the vaccinees consistently clustered into two groups based on the level of their NAb titer: low and high (Figure 5A and 5B). Using three EC50 thresholds (10, 20, 30), high-titer NAb group had a lower number of escape mutations compared to low-titer NAb group (Figure 5C).

**Figure 5.**
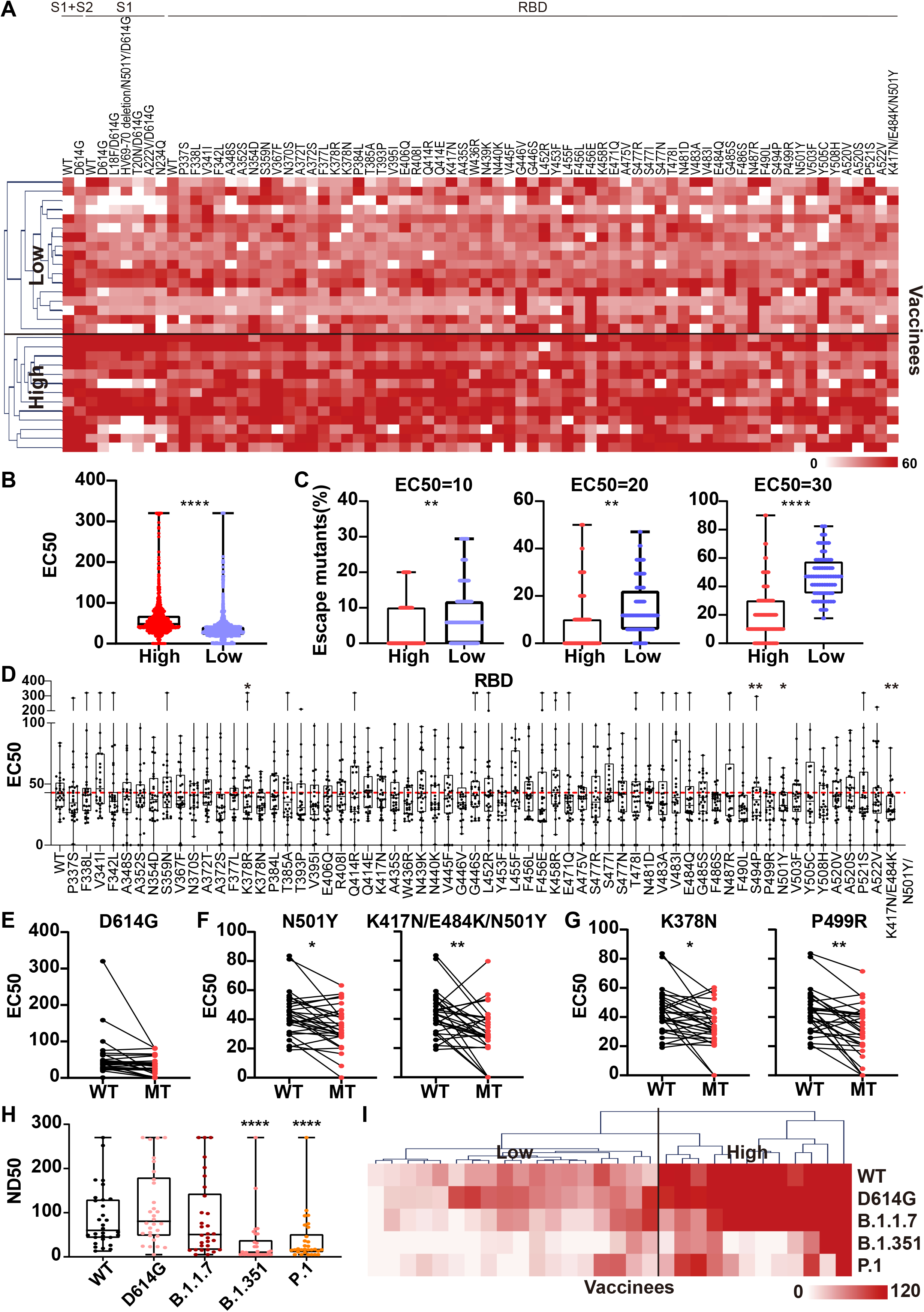
Heterogeneous responses of serological NAbs to the SARS-CoV-2 S mutations in vaccinated individuals. (A) Hierarchical cluster analysis of NAbs to S variants in vaccinees. The rainbow color from white to red correspond the NAb titers from low to high, respectively. (B) EC50 comparison of vaccinees with high and low NAb titers. (C) Comparison of escape mutations between vaccinees with high and low NAb titers with EC50 thresholds set at 10, 20 and 30, respectively. (D) Distribution of serological NAbs to different SARS-CoV-2 S variants in vaccinees. (E) Comparison of NAbs between the wild-type and D614G S proteins. (F) Comparison of NAbs between the wild-type S protein and S variants with known escape mutations. (G) Comparison of NAbs between the wild-type S protein and S variants with newly-identified escape mutations. Significant escape mutations were identified using an unpaired t test with a p-value of 0.05. (H) Detection of serological NAbs to prevalent SARS-CoV-2 variants in the vaccinees using the pseudovirus neutralization assay. (I) Hierarchical cluster analysis of the NAbs to prevalent SARS-CoV-2 variants in vaccinees. The rainbow color from white to red correspond the NAb titers from low to high, respectively.

Next, the NAb titers of vaccinees across all S variants were ascertained (Figure 5D). Like the convalescent COVID-19 patients, the D614G variant did not alter the neutralizing effect of the NAbs compared to wild-type S1+S2, S2, and RBD (Figure 5E, Figure S9). Using statistical analysis, four escape mutations were identified, including 2 known (N501Y, K417N/E484K/N501Y) and 2 new (K378N, P499R) mutations (Figure 5F and 5G) (Chen et al., 2021).

To validate the results obtained with the mSAIS assay, we analyzed the NAbs with a SARS-CoV-2 pseudovirus neutralization assay displaying wild-type and mutant S (wild-type, D614G, B.1.1.7, B.1.351, P.1). The results for the D614G, B.1.1.7, B.1.351 variants aligned with the data obtained with the mSAIS assay (Figure 5H). The vaccinees also separated into high- and low-titer NAb groups (Figure 5I), further supporting the data obtained with the mSAIS assay Figure 5C).

### High titer of NAbs contain anti-Spike antibodies with diverse binding epitopes

Two recent studies with rhesus macaques have shown that a high titer of NAbs produced in response to vaccination (DNA-based or adenovirus serotype 26 (Ad26) vector-based) provided good protection against infection when challenged with SARS-CoV-2 (Mercado et al., 2020; Yu et al., 2020). In another study, NAb titers were significantly higher in vaccinees who had COVID-19 previously than vaccinees who never had COVID-19. Moreover, the vaccinees who had been infected with SARS-CoV-2 possessed NAbs that were able to neutralize all prevalent S variants (Stamatatos et al., 2021).

In order to provide insight into how well the high-titer NAb samples could protect against SARS-CoV-2 variants, we analyzed the differential expression of anti-S antibodies in the vaccinees. The level of antibodies that target the S protein’s S1, S2 extracellular domain (ECD), and RBD is higher in the high-titer NAb group than the low-titer NAb group. Of these three antibodies, the differing level of anti-RBD antibodies across the two groups was the most pronounced (Figure 6A).

**Figure 6.**
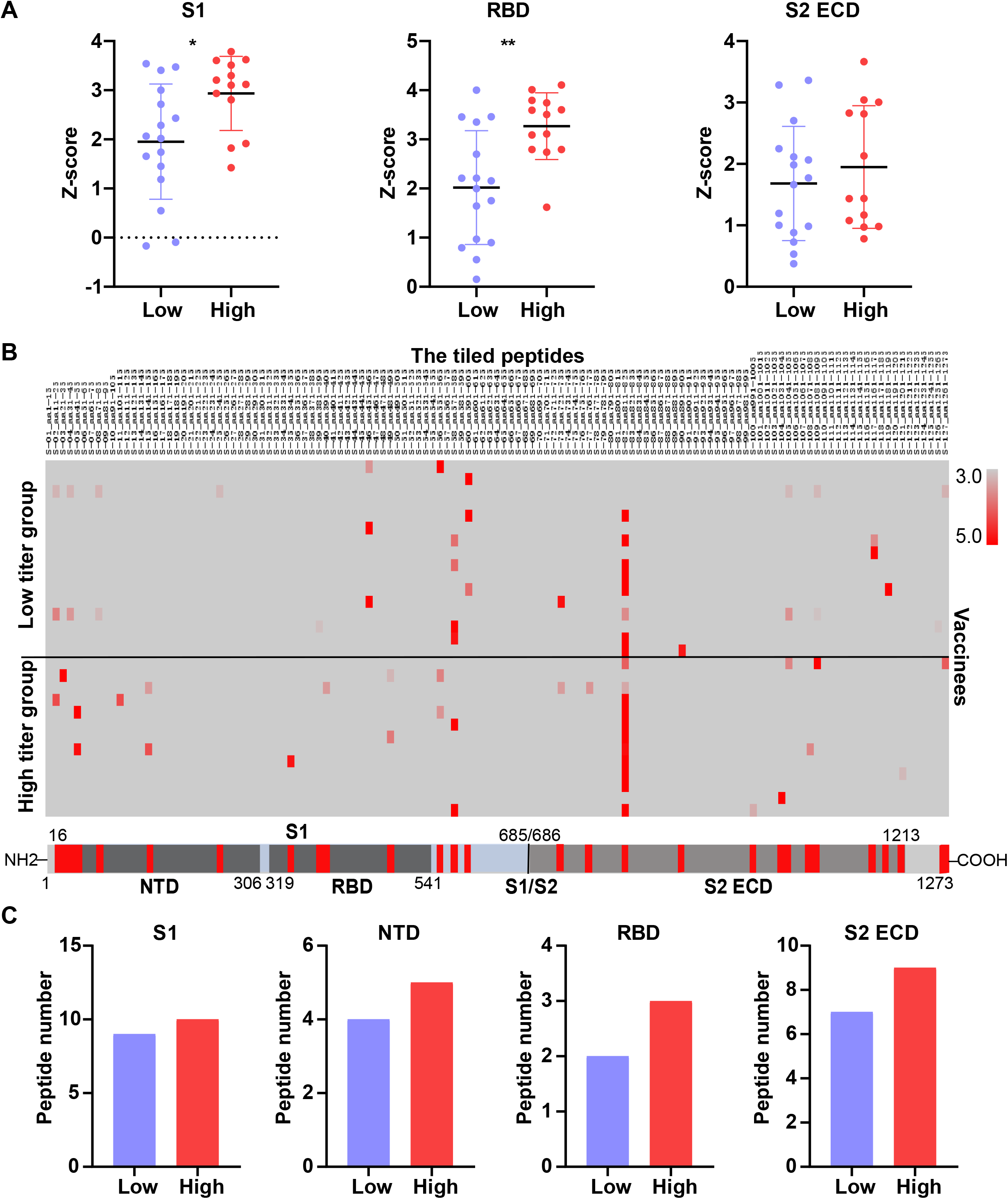
The NAb titer is associated with the diversity of anti-S antibodies in the serum of vaccinees. (A) A comparison of levels of antibodies that target the S1, RBD and S2 ECD. (B) Identification of vaccine-induced humoral immune responses to the S protein. (C) Association between the number of immunogenic peptides and NAb titers. A p-value (p) < 0.05 by Mann-Whitney test was considered to be significant. NTD = N-terminal domain

Next, we performed epitope mapping of the vaccinees’ serological anti-S antibodies using a tiled peptide microarray representing the full-length S protein as previously described (Liang et al., 2021; Wang et al., 2020). Indeed, the results indicate that the high-titer NAb group bound to more diverse epitopes than the low-titer NAb group (Figure 6B). The number of immunogenic peptides within the S1, S2ECD, and RBD is also consistently higher in the high-titer NAb group (Figure 6C).

To further validate our findings, sera from the 104 vaccinees were analyzed using a SARS-CoV-2 pseudovirus neutralization assay. Prior to data analysis, all vaccinees were separated into three groups according to their NAb titers (<10, 10-50, >50). Notably, the results show that the concentration of anti-S antibodies is proportional to the concentration of NAbs. However, there was no difference in the levels of anti-S1 antibodies that target the N-terminus or C-terminus across the vaccinees. The levels of anti-S2 ECD antibodies were significantly different only between NAb titers of <10 and >50 (Figure 7A). These results demonstrate that the antibodies produced in response to vaccination of inactivated vaccine primarily target the RBD (Figure 6A and 7A).

**Figure 7.**
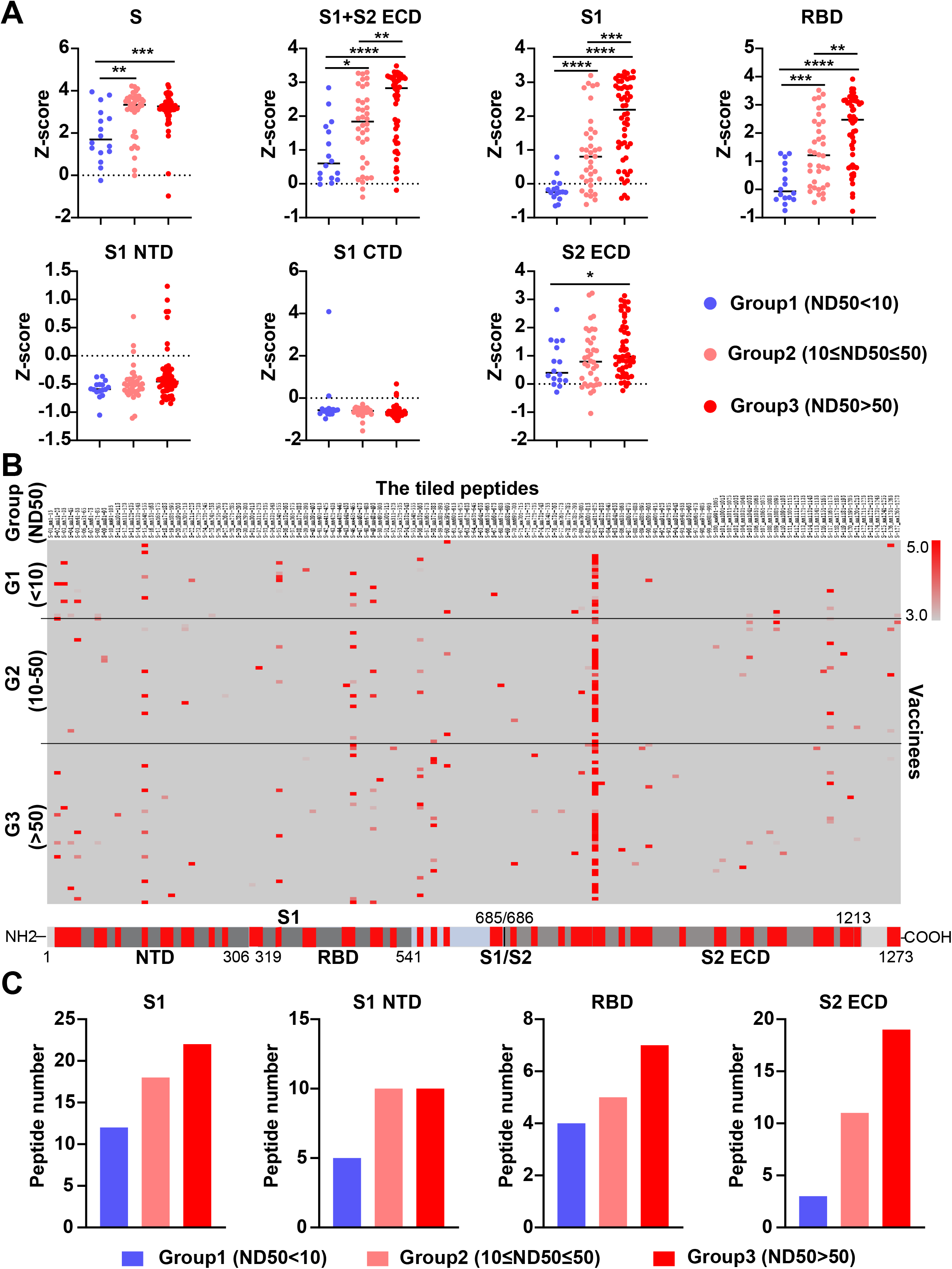
Validation of anti-S antibody diversity in 104 vaccinees. (A) A comparison of levels of antibodies that target the S proteins and its domains. (B) Landscape of vaccine-induced humoral immune responses to the S protein. (C) Association between the number of immunogenic peptides and NAb titers. A p-value (p) < 0.05 by Mann-Whitney test was considered to be significant. CTD = C-terminal domain; NTD = N-terminal domain. The NAb titers (ND50) was measured using pseudovirus neutralization assay.

The peptide microarray was also employed to examine serological NAbs with a larger vaccinated cohort (n=104). The data confirm that the anti-S antibodies in the high-titer NAb group bind to more peptides representing the S1, S1 N-terminal domain, RBD, and S2 ECD of the S protein than the low-titer NAb group (Figure 7B, 7C). These data indicate that high NAb titers contain anti-S antibodies with increased binding diversity.

## Discussion

The SARS-CoV-2 virus has mutated over time, resulting in circulating viral strains with altered transmission efficiencies, mortality rates, and S variants that can escape from the neutralizing effect of antibodies (Alam et al., 2021) (Davies et al., 2021; Hou et al., 2020b; Prevost and Finzi, 2021). Therefore, significant effort has been devoted to improve the diagnosis, prevention, and treatment of COVID-19 patients infected with SARS-CoV-2 variants (Abdool Karim and de Oliveira, 2021).

In this work, we developed an mSAIS assay that enables the detection of NAbs to numerous S variants simultaneously and rapidly. It has numerous advantages compared to the enzyme-linked immunosorbent assay (ELISA) and pseudovirus neutralization assays (Li et al., 2021; Tan et al., 2020; Weisblum et al., 2020). First, our flexible platform can be easily adapted to include new S variants as they emerge. Second, only a laser scanner capable of detecting Cy5 is required to perform the mSAIS assay. Third, the multiplexed nature of the platform significantly reduces the sample volume, reagents, time, and cost to obtain data compared to single-plexed assays (Xu et al., 2020). For example, only 20 µL of serum was needed to screen for NAbs against 72 S variants in this study. In comparison, the volume requirements for ELISA or the pseudovirus neutralization assay would be 1,440 (72×) or 3,240 (162×) µL, respectively.

Using the mSAIS assay, we compared purified mouse and rabbit antibodies. Of the three antibodies tested, antibody #53 was able to inhibit the S-ACE2 interaction across the most S variants, thus it is a superior candidate for COVID-19 therapy. The comprehensive mapping of antibodies to different S variants would be valuable in understanding the sensitivity of mutations to antibody neutralization, and help develop antibody cocktails for COVID-19 therapy (Li et al., 2021; Li et al., 2020; Prevost and Finzi, 2021; Starr et al., 2021).

Using serum from convalescent COVID-19 patients and vaccinees with the mSAIS assay, we identified 4 and 2 new resistant mutations, respectively, that escape NAb recognition (Figures 4 and 5). We also show that there are fewer escape mutations in high NAb titers than low titers, and this was observed in both convalescent COVID-19 patients and vaccinees (Figures 4C and 5C). Using a large cohort of 104 vaccinees with multiple assays (i.e., mSAIS assay, peptide array, pseudovirus neutralization assay), we further demonstrate that high titer NAbs contain anti-S antibodies that target more diverse binding epitopes, thus leading to more neutralizing capacity across a breadth of S variants (Figure 6 and 7). The production of heterogenous antibodies could be due to the somatic mutations that occur during antibody maturation (Muecksch et al., 2021). Finally, our data suggest that an effective COVID-19 vaccination strategy to defend against the mutating SARS-CoV-2 is to elicit the immune response to produce high-titer NAbs that contain a large diversity of anti-S antibodies (Stamatatos et al., 2021; Starr. et al., 2021). However, the association between NAb titer and its ability to protect against SARS-CoV-2 infection in humans is unclear (Jin et al., 2021).

There were several limitations in this study. First, the mSAIS assay is *in vitro*, which might not accurately reflect results *in vivo*. Second, some conformational and glycosylation epitopes might not be detected using the peptide microarray that employed chemically-synthesized peptides (Wang et al., 2020). Finally, the number of serum samples from convalescent COVID-19 patients and vaccinees were limited. The results obtained in this study should be validated in a large different cohort in the future.

## Conclusion

Altogether, we developed a high-throughput microarray-based mSAIS assay, which enables the multiplexed and rapid screening of NAbs to SARS-CoV-2 S variants. Using the mSAIS assay, we confirmed the neutralization capabilities of NAbs to known mutations, identified a number of new mutations that are resistant to the NAbs, and showed that there were fewer escape mutations with high NAb titers than low NAb titers in both convalescent COVID-19 patients and vaccinees. The data demonstrate the great potential of our proteomics platform in mapping the ability of NAbs to block the interaction between SARS-CoV-2 S variants and ACE2 interactions. Although we tested purified antibodies and serum in this study, the mSAIS assay could be used with other sample types as well. Data gleaned from the mSAIS will help develop more effective vaccines and therapeutic antibodies to fight against COVID-19.

## Data Availability

The ability of the serological NAbs to inhibit the S-ACE2 interactions across the different S variants was visualized as a heatmap using the MultiExperiment Viewer software version 4.9 (Chu et al., 2008). Statistical analyses were performed using the GraphPad Prism software 8.3 and Microsoft Excel with the unpaired t test and Mann-Whitney test. A p-value (p) < 0.05 was considered to be significant. *p < 0.05, **p < 0.01, ***p < 0.005, ****p < 0.001. The 3D structure of the SARS-CoV-2 RBD and ACE2 complex (PDB ID code: 6M0J) was visualized using the VMD 1.9.3, and mutations were annotated.

## Contributions

J.Z. and L.X. provided the purified spike variant proteins. M. Z., S. Y., C. H., Y. H., and Y. W. provided the clinical samples. X. Z., H. W., J. Z., J. R., H. B., and X. Y. executed microarray experiments. X. Z., T. L., H. P., S. L., C.W., and X.Y. executed the bioinformatics and statistical analyses. J.V. executed the pseudovirus based neutralization assay. H. Z., M. L., X. H., and B. K. executed the authentic virus-based neutralization assay. X.Y., Y. W., and B. K. conceived the idea, designed experiments, analyzed the data, and wrote the manuscript.

## Acknowledgement

This work was supported by the National Natural Science Foundation of China (31870823), National Key R&D Program of China (2020YFE0202200), State Key Laboratory of Proteomics (SKLP-C202001, SKLP-O201904, SKLP-O201703, SKLP-O202007), the National Program on Key Basic Research Project (2018YFA0507503, 2017YFC0906703, 2018ZX09733003) and the Beijing Municipal Education Commission, Beijing Municipal Natural Science Foundation (M21003) and Beijing Municipal Science&Technology Commission (Z201100001020001; Z201100005420022). We thank Dr. Brianne Petritis for her critical review and editing of this manuscript.

## Supplemental information

The supplemental information includes 3 supplementary tables and 9 supplementary figures.

## Materials and methods

### Collection of patient samples

Clinical specimens were obtained from the Department of Clinical Laboratory in Beijing Ditan Hospital, Capital Medical University (Beijing, China). Whole blood was collected in a vacutainer tube, and centrifuged at 4,000×g at room temperature (RT) for 10 minutes to separate serum. Serum was then transferred to a clean tube and stored at -80°C until use. For safety purposes, serum collected from convalescent COVID-19 patients was inactivated at 56°C for 30 minutes prior to any further sample processing. Notably, serum from COVID-19 vaccinated individuals was obtained from healthy hospital staff 4 weeks after receiving the second vaccine dose, and convalescent samples were collected from COVID-19 patients 2 to 4 weeks post discharge. Our research was approved by the Ethics Committee of Beijing Ditan Hospital (No. 2021-010-01), and exemption of informed consent was obtained prior to sera collection.

### Expression and purification of recombinant proteins for use on the mSAIS assay

The DNA sequence encoding the SARS-CoV-2 Spike protein (S1+S2 ECD) (YP_009724390.1) (Val 16-Pro1213), S1 Subunit (YP_009724390.1) (Val16-Arg685), RBD (YP_009724390.1) (Arg319-Phe541) or variants were expressed with a polyhistidine tag at the C-terminus. The generation of variants using site-directed mutagenesis was performed as previously described (Li et al., 2020). In addition, two negative controls [SARS-CoV-2 Nucleocapsid (N), MERS-CoV-2 Spike RBD], one positive control (SARS-CoV-2 wild-type RBD), and the extracellular domain of ACE2 were expressed with a polyhistidine tag at the C-terminus. The proteins were expressed in HEK293 cells, followed by further purification on a Ni-NTA spin column (Thermo Fisher). Protein purity was confirmed using an SDS-PAGE gel stained with Coomassie blue.

### Preparation of the mSAIS microarray

All purified proteins were diluted to 100 µg/mL and printed onto a three-dimensional (3D)-modified slide surface (Capital Biochip Corp, Beijing, China) in parallel and in duplicate using an Arrayjet microarrayer (Roslin, UK) as previously described (Hou et al., 2020a; Wang et al., 2020; Xu et al., 2019). The protein microarrays were stored at -20 °C until ready to use.

### Detection of NAbs using the mSAIS assay

Prior to antibody detection, the protein microarrays were assembled in an incubation tray and blocked with 1% (w/v) milk in 1x phosphate buffered saline, pH 7.2 (PBS), with 0.2% (v/v) Tween-20 (PBST) for 10 min at room temperature. After washing with PBST three times, the array was incubated with serum diluted 10-to 320-fold in PBST for 30 min at room temperature. After washing again, the array was then incubated for 60 min with Cy5-labeled ACE2 (50 ng/mL). Finally, the array was washed with PBST and water, dissembled from the tray, and dried via centrifugation for 2 min at 2,000 rpm. The array was scanned with a GenePix 4300A microarray scanner (Molecular Devices, Sunnyvale, CA, USA) at a 10 μm resolution using a laser at 635 nm with 100% power/ PMT Gain 900. The median fluorescent signal intensity with background subtraction was extracted using GenePix Pro7 software (Molecular Devices, Sunnyvale, CA, USA).

### SARS-CoV-2 pseudovirus neutralization assay

The SARS-CoV-2 pseudovirus neutralization assay was performed as previously described (Nie et al., 2020). Briefly, human sera were diluted using 3-fold serial dilutions with a starting concentration of 1:10 (two per dilution). The diluted sera were added into 96-well plates in duplicate, followed by the SARS-CoV-2 pseudovirus at a concentration of 1300 median tissue culture infective dose (TCID50/mL). The sera and pseudovirus were incubated together at 37°C for 1 hour, and then monkey Vero cells were added at 2*10^4 cells/100 mL cells per well. The plates were incubated at 37°C in a humidified atmosphere with 5% CO_2_ for 24 hours before chemiluminescence detection was performed. The Reed-Muench method was used to calculate the neutralization titer.

### Live SARS-CoV-2 neutralization assay

A suspension of Vero cells suspension was prepared at a cell density of 2×10^5^/ml. Serum samples were diluted 4-fold in cell culture medium and inactivated at 56°C for 30 minutes. The inactivated serum was serially diluted 2-fold with MEM medium, followed by the addition of cell culture medium containing 100 times the cell culture infective dose 50% (CCID50) of wild-type SARS-CoV-2 virus. The mixture was placed into a 96-well plate and incubated at 37°C for 2 h in CO_2_ 5%, and then 2□×□10^4^ Vero cells were added and incubated again at 37°C. The samples were microscopically examined for cytopathic effect (CPE) after 7 days. The highest dilution of serum that showed complete inhibition activity of SARS-CoV-2 was recorded as the neutralizing antibody titer (ID50). Assays were performed in duplicate with negative control serum.

The ID50 was calculated as: LogCCID50 = Xm + 1/2d - d ∑pi/100, where Xm is the log10 of maximum dilution of the virus, the d is the logarithm of the dilution fold, and ∑pi is the sum of the percentage of the cytopathic effect per dilution.

### Detection of SARS-CoV-2 serum antibodies using a SARS-CoV-2 proteome peptide microarray

The peptide microarray containing truncated S proteins, and tiled peptides representing SARS-CoV-2 S protein was prepared as our previous work (Liang et al., 2021; Wang. et al., 2020). The array was assembled in an incubation tray and blocked with 5% (w/v) milk in phosphate-buffered saline (PBS) containing 0.05% (v/v) Tween-20 (PBST) for 30 min at room temperature before antibody detection. After aspirating the blocking solution, the 1:200 diluted serum was added to the array and incubated at room temperature for 20 min. After washing three times with PBST, the array was then incubated for 20 min with a Cy3 labelled donkey anti-human IgG(H+L) antibody (Jackson ImmunoResearch, USA) (2 μg/mL). Finally, the array was washed with PBST and deionized water, disassembled from the tray and dried with vacuum pump. The slide was scanned at 532 nm using a GenePix 4300A microarray scanner (Molecular Devices, Sunnyvale, CA, USA). The median fluorescent signal intensity of each spot with background subtraction was extracted using GenePix Pro7 software (Molecular Devices, Sunnyvale, CA, USA).

