## Supplementary Text and Figures for "Inhibitor screening of Spike variants reveals the heterogeneity of neutralizing antibodies to COVID-19 infection and vaccination"

**Supplementary tables**

| Table S1 | Clinical information of convalescent COVID-19 patients. |
| --- | --- |
| Table S2 | Clinical information of people received with COVID-19 inactivated vaccine. |
| Table S3 | Clinical information of validation cohort comprising of people received with COVID-19 inactivated vaccine. |

**Supplementary figures**

| Figure S1 | SDS-PAGE analysis of purified recombinant SARS-CoV-2 S variants. |
| --- | --- |
| Figure S2 | Timeline of when the top 10 RBD mutations emerged. |
| Figure S3 | Layout of SARS-CoV-2 spike variant protein microarray. |
| Figure S4 | Reproducibility of protein microarray preparation. |
| Figure S5 | Correlation of mSAIS assay and live SARS-CoV-2 neutralization assay. |
| Figure S6 | Detection of different NAbs binding to immobilized RBD protein using the mSAIS assay. |
| Figure S7 | Structural analysis of spike mutations that change the neutralization activities of purified antibodies. |
| Figure S8 | NAb titers to the wild type and variant S proteins in convalescent COVID-19 patients. |
| Figure S9 | NAb titers to the wild type and variant S proteins in vaccinees. |

**Supplementary tables**

**Table S1: Clinical information of the convalescent COVID-19 patients.**

| Gender | n | Age, median | Weeks post discharge |
| --- | --- | --- | --- |
| Female | 11 | 49 (18 ~ 64) | 2 or 4 |
| Male | 14 | 36 (5 ~ 68) | 2 or 4 |
| Total | 25 | 43 (5 ~ 68) | 2 or 4 |

**Table S2: Clinical information of people received with COVID-19 inactivated vaccine.**

| Gender | n | Age, median | Weeks post the  second vaccine dose |
| --- | --- | --- | --- |
| Female | 6 | 36 (21 ~ 53) | 4 |
| Male | 24 | 37 (23 ~ 57) | 4 |
| Total | 30 | 37 (21 ~ 57) | 4 |

**Table S3： Clinical information of validation cohort comprising of people received with COVID-19 inactivated vaccine.**

| Gender | n | Age, median | Weeks post the second vaccine dose |
| --- | --- | --- | --- |
| Female | 11 | 46 (21 ~ 70) | 4 |
| Male | 93 | 39 (23 ~ 70) | 4 |
| Total | 104 | 39 (21 ~ 70) | 4 |

**Supplementary Figures**

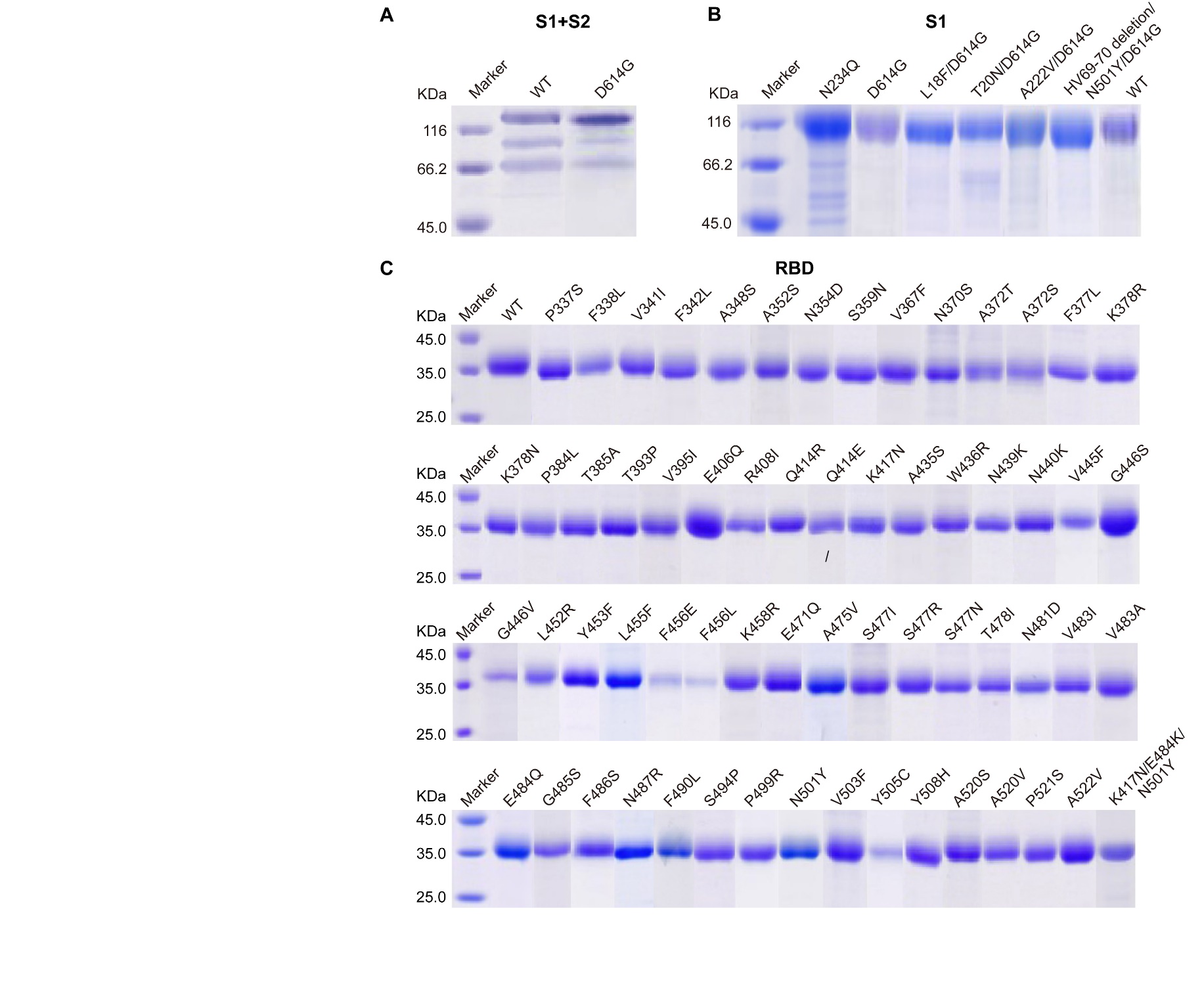

**Figure S1. SDS-PAGE analysis of purified recombinant SARS-CoV-2 S variants.** (A-C) are the S1+S2, S1 and RBD proteins, respectively.

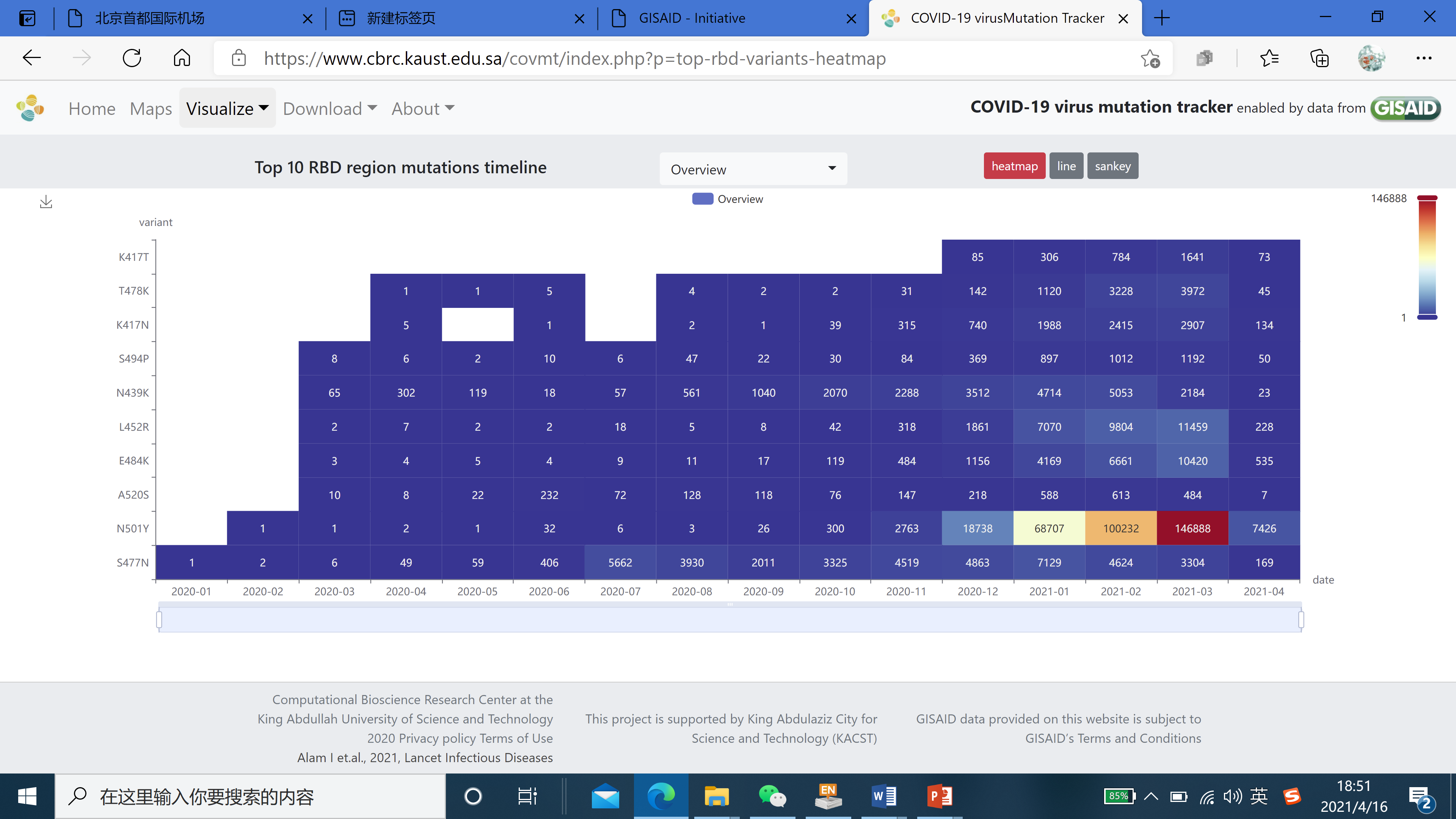

**Figure S2. Timeline of when the top 10 RBD mutations emerged.** The graph was obtained from the COVID-19 virus mutation tracker database (https://www.cbrc.kaust.edu.sa/covmt/index.php?p=top-rbd-variants-heatmap)(Alam et al., 2021).

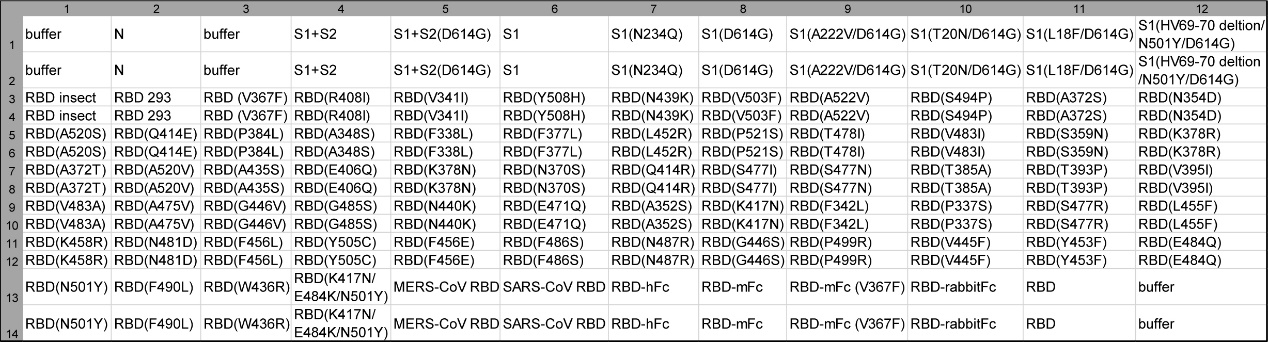

**Figure S3. Layout of SARS-CoV-2 spike variant protein microarray.** The buffer and nucleocapsid (N) protein served as the negative controls. The RBD protein served as the positive control.

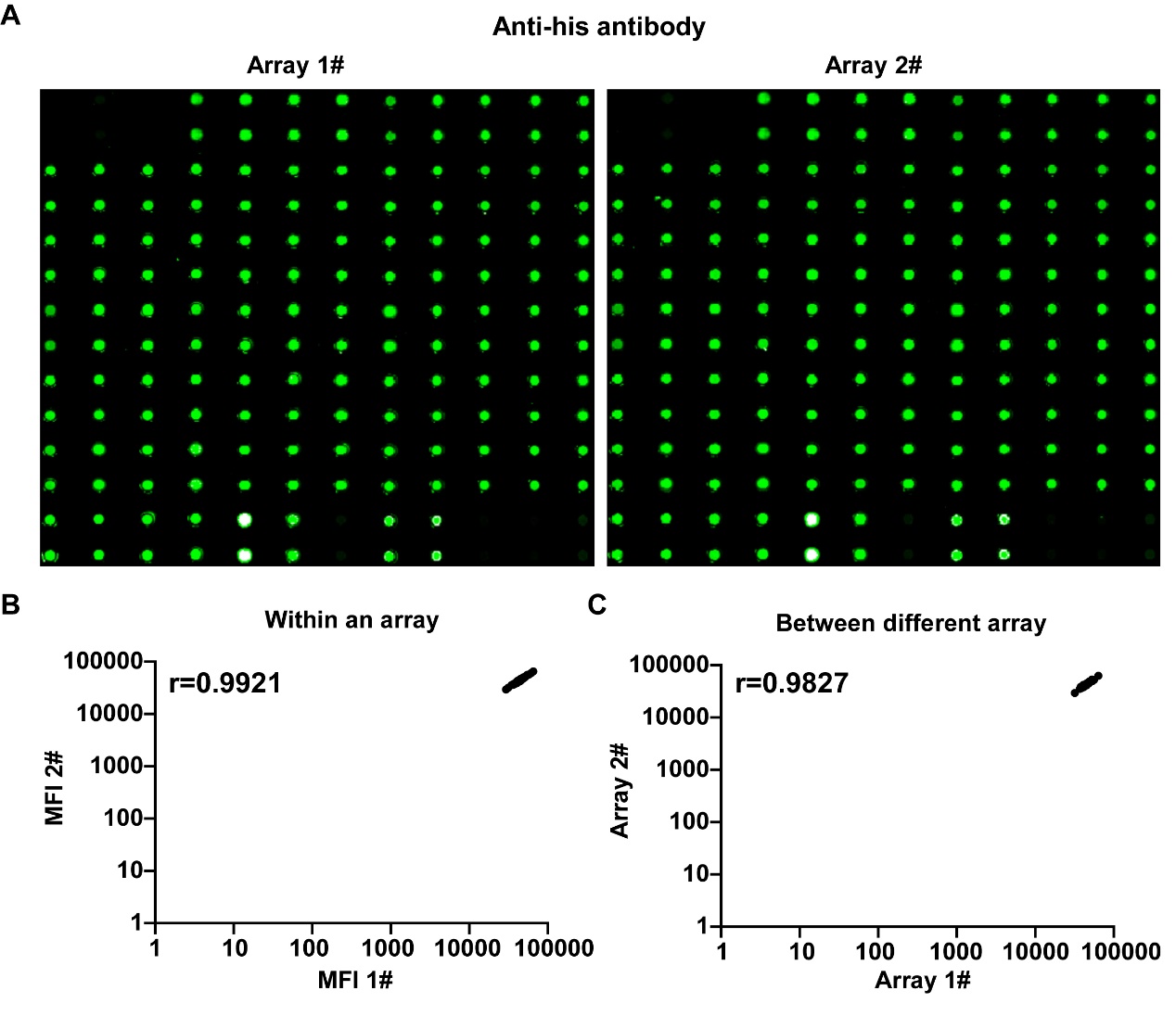

**Figure S4. Reproducibility of protein microarray preparation.** (A) Fluorescent staining of immobilized proteins on the glass-based microarray using an anti-his antibody; (B-C) are the intra- and inter-array reproducibility, respectively.

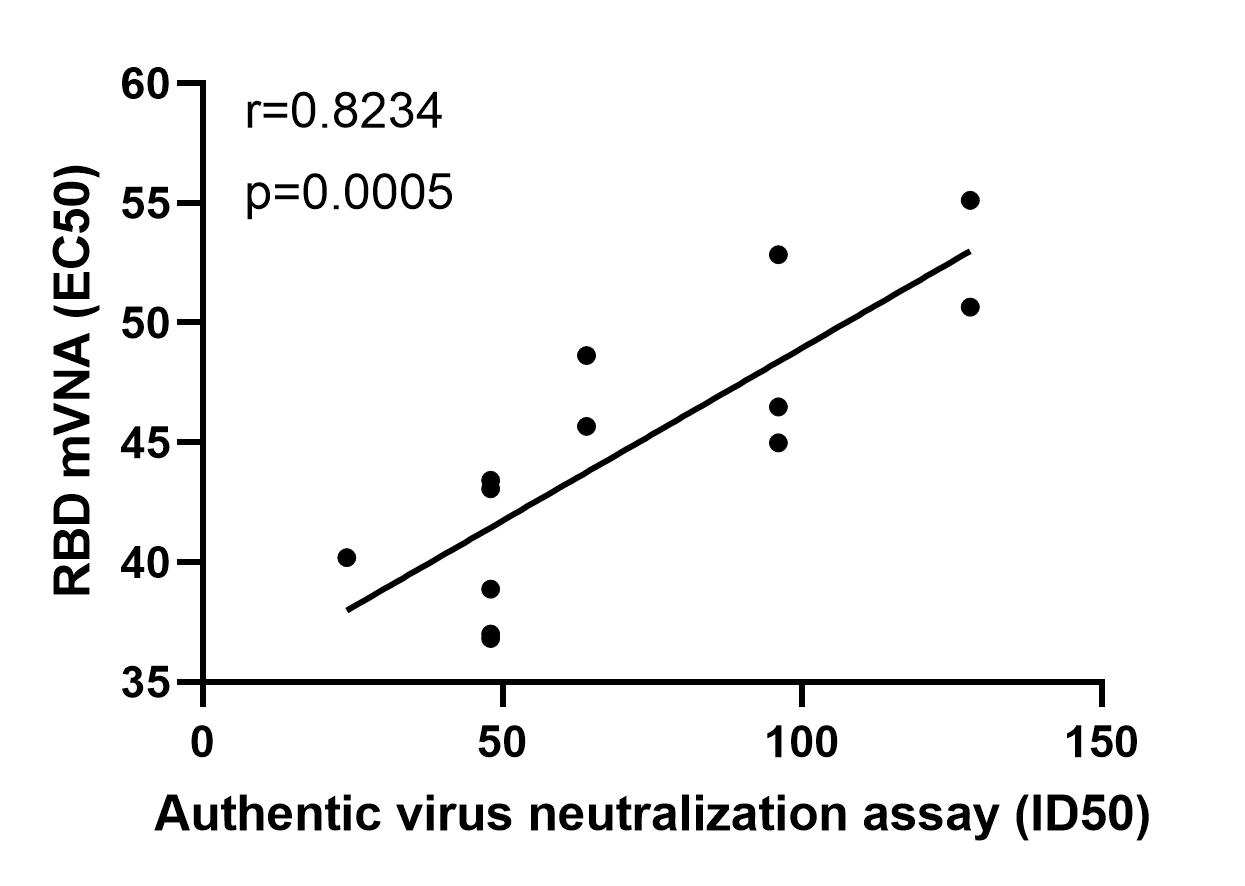

**Figure S5. Correlation of mSAIS assay and live SARS-CoV-2 neutralization assay.** Pearson’s correlation coefficient and linear regression analyses were performed by the GraphPad Prism software 8.3. Statistical significance was determined using the two-tailed t-test.

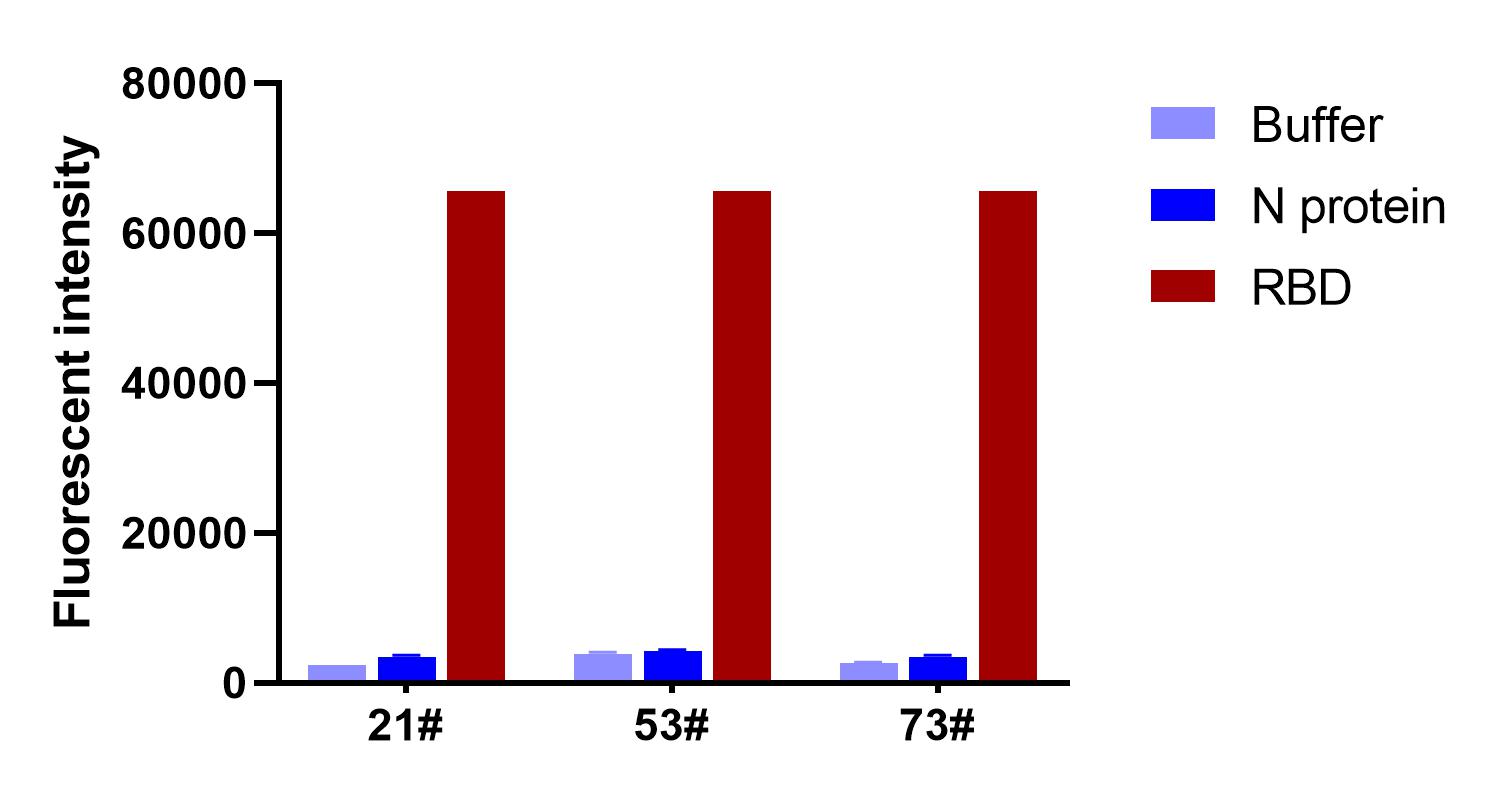

**Figure S6. Detection of different NAbs binding to immobilized RBD protein using the mSAIS assay.** The nucleocapsid (N) protein and buffer served as the negative controls.

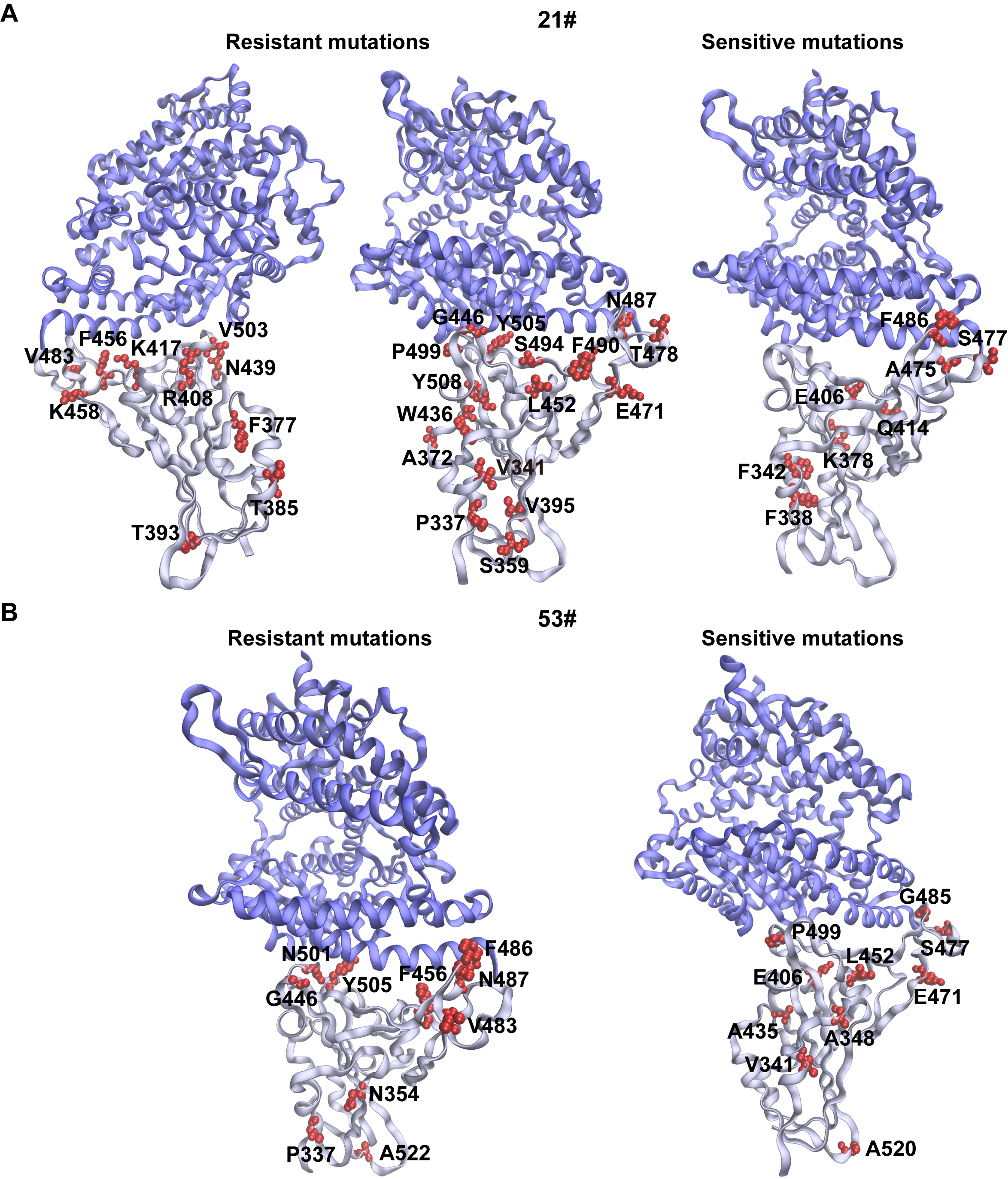

**Figure S7. Structural analysis of spike mutations that change the neutralization activities of purified antibodies.** (A, B) The structural analyses of the mutations that are resistant and sensitive to antibbodies #21 and #53, respectively. The interaction structure of the SARS-CoV-2 RBD and human ACE2 were derived from the Protein Data Bank with PDB ID 6M0J.

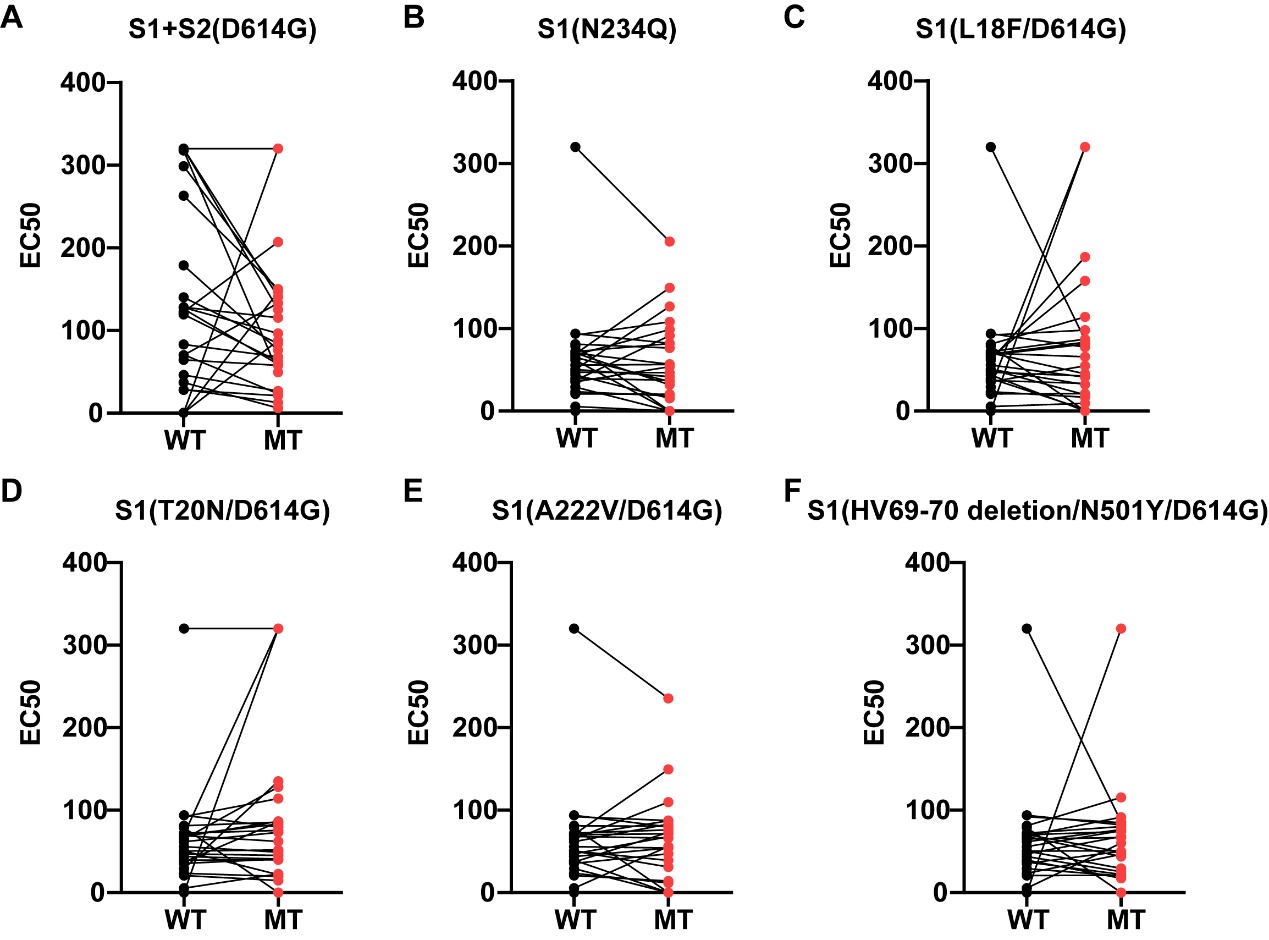

**Figure S8. NAb titers to the wild type and variant S proteins in convalescent COVID-19 patients.** (A-F) are the testing of different S variants with D614G, N234Q, L18F/D614G, T20N/D614G, A222V/D614G and hv69-70 deletion/N501Y/D614G, respectively. WT and MT represent the wild type and mutants, respectively.

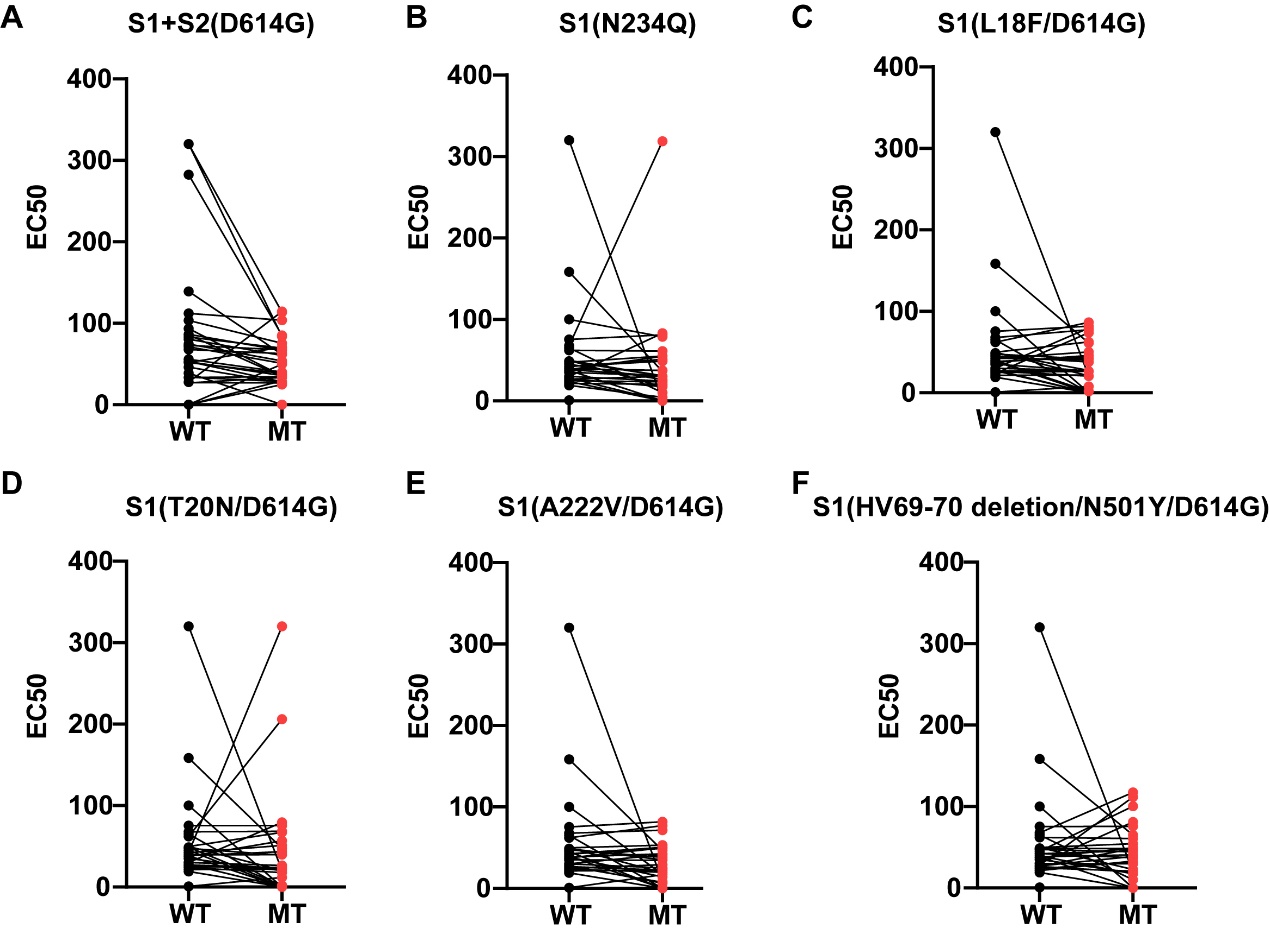

**Figure S9. NAb titers to the wild type and variant S proteins in vaccinees.** (A-F) are the testing of different S variants with D614G, N234Q, L18F/D614G, T20N/D614G, A222V/D614G and hv69-70 deletion/N501Y/D614G, respectively. WT and MT represent the wild type and mutants, respectively.
